# Maternal and Neonatal Outcomes Associated with Prenatal Prescription Stimulant Treatment Among Pregnant Individuals with Opioid Use Disorder

**DOI:** 10.64898/2026.11.12.26367944

**Authors:** Thomas J. Reese, Ashley A. Leech, Julia C. Phillippi, Stephen W. Patrick, Elizabeth A McNeer, Sarah Loch, Jessica Young, Amanda von Horn, Mauli V. Shah, Andrew D. Wiese

**Author notes:** **Corresponding Author** Thomas J. Reese, PharmD, PhD, Vanderbilt University Medical Center, Department of Biomedical Informatics, 2525 West End Avenue, Suite 1475.

## Abstract

**Introduction:** Clinicians and Medicaid programs must decide whether to continue prescription stimulants in pregnancies complicated by opioid use disorder (OUD), yet no safety evidence exists for this population and stimulant coverage is frequently restricted. We evaluated maternal and neonatal outcomes associated with prescription stimulant treatment during pregnancy among individuals with OUD, including those with co-occurring stimulant use disorder (StUD) and attention-deficit/hyperactivity disorder (ADHD)

**Methods:** We conducted a retrospective cohort study of Medicaid⍰insured pregnant individuals in Tennessee (2010–2021) with OUD. Our exposure of interest was any prescription amphetamine⍰ or methylphenidate⍰class stimulant overlapping pregnancy. The primary maternal outcome was overdose from delivery through 1 year postpartum; secondary outcomes included death, severe maternal morbidity and cardiovascular complications in the first 6 weeks postpartum. The primary neonatal outcome was preterm birth; secondary outcomes included low birth weight, NICU admission, neonatal opioid withdrawal syndrome (NOWS), and death. We used overlap weighting to estimate adjusted odds ratios (aORs) and predicted probabilities and evaluated effect modification by buprenorphine.

**Results:** Among 12,327 pregnancies with OUD, 380 (3.1%) received prescription stimulants during pregnancy; stimulant treated individuals were slightly older and more likely to have mental health comorbidities. In adjusted analyses, stimulant treatment was not associated with postpartum overdose (aOR, 1.13; 95% CI, 0.61–2.08), preterm birth (aOR, 1.04; 95% CI, 0.74–1.46), cardiovascular complications, severe maternal morbidity, low birth weight, or NOWS. Odds of NICU admission were higher among stimulant exposed infants (aOR, 1.58; 95% CI, 1.17–2.14). Maternal and infant deaths were rare. In exploratory analyses, overdose odds were elevated among pregnancies receiving both stimulants and buprenorphine (P for interaction = .03) and the cardiovascular composite was elevated within the ADHD subgroup (aOR, 2.33; 95% CI, 1.13–4.82).

**Conclusions:** These first OUD-specific estimates do not support routine discontinuation of clinically indicated stimulant therapy in pregnancy. Higher NICU admission among stimulant-exposed infants, unaccompanied by differences in preterm birth, low birth weight, gestational age, or NOWS, is more consistent with intensified surveillance than with neonatal harm. Two exploratory signals require replication before they can inform practice.

## INTRODUCTION

Drug overdose is a leading cause of pregnancy-associated mortality in the United States, increasingly driven by polysubstance use involving opioids and stimulants.^1–3^ Pregnant individuals with opioid use disorder (OUD) have well-documented increased risks of maternal overdose, infection, severe maternal morbidity, and death.^1–3^ Infants born to individuals with OUD also face increased risks of preterm birth, low birth weight, neonatal intensive care unit (NICU) utilization, and neonatal opioid withdrawal syndrome (NOWS).^4–6^ Similarly, pregnant individuals with stimulant use disorder (StUD) have increased risks of poor maternal and infant outcomes, including hepatitis and human immunodeficiency virus infection, vasoconstrictive cardiovascular complications, fetal growth restriction, and preterm birth.^7–12^ Yet, despite the growing involvement of stimulants in overdose deaths and the increasing prevalence of perinatal polysubstance exposure, the effect of co-occurring StUD and OUD during pregnancy is not well understood.^1,2^

Prescription stimulants, including amphetamine- and methylphenidate-class medications, are commonly prescribed for ADHD and may also be used off-label among individuals with co-occurring OUD and StUD.^12,13^ Population-based studies of pregnant individuals suggest that prescription stimulants alone are not associated with major congenital anomalies or increased neurodevelopmental risk, and some observed increased risks are partially explained by confounding by indication.^13–18^ Given the documented harms of untreated ADHD and StUD in pregnancy, current recommendations emphasize shared decision-making that weighs stimulants risks against risks of inadequately treated disease.^12,19^ For obstetric clinicians, this decision often arises at entry to prenatal care, yet safety data specific to pregnancies complicated by OUD are limited. Coverage policy compounds the uncertainty: state Medicaid programs commonly apply prior authorization, quantity limits, or outright exclusions to stimulants, so decisions to continue or discontinue treatment are frequently made by payers and prescribers without population-specific evidence. Emerging evidence further suggests prescription stimulants may reduce illicit stimulant use in some contexts, although evidence remains limited and heterogeneous.^7–9^

Our objective was to estimate the association between prenatal prescription stimulant treatment and maternal and neonatal outcomes among individuals with OUD overall and stratified by indication for stimulant use (ADHD and StUD). Because this decision is already being made in routine practice without OUD-specific evidence, we framed the analysis as a safety assessment rather than a test of a directional hypothesis, and prespecified that estimates would be interpreted on the basis of the magnitude of risk compatible with the confidence limits, not statistical significance alone.

## METHODS

### Study Design and Data Sources

We conducted a retrospective cohort study of pregnant individuals enrolled in Tennessee Medicaid (TennCare). TennCare administrative claims were linked to Tennessee birth and death certificates from the Tennessee Department of Health for deliveries occurring January 1, 2010, through December 31, 2021. Pregnancy was defined from the estimated last menstrual period (LMP) through the delivery hospitalization. The study was approved by the Vanderbilt University Medical Center and Tennessee Department of Health institutional review boards and the Division of TennCare, with data use agreements in place, and followed the STROBE reporting guideline for cohort studies.^20^

### Study Population and Eligibility Criteria

We identified all deliveries during the study period and restricted to pregnancies among individuals with OUD, defined as evidence of diagnosis from at least 1 inpatient or 2 outpatient maternal encounter claims separated by at least 7 days using *International Classification of Diseases, Ninth and Tenth Revision, Clinical Modification* (ICD-9-CM and ICD-10-CM) codes.^6^ We included pregnancies among individuals aged 15 to 44 years at delivery, singleton gestation, and for the primary analysis examining overdose required continuous TennCare enrollment in the 180 days before delivery through 365 days postpartum. For secondary analyses we required continuous enrollment in the 180 days before delivery through 42 days postpartum for cardiovascular complications and severe maternal morbidity. For neonatal outcomes, we required infant enrollment from birth through 30 days after birth. Because eligibility requirements after delivery may limit capture of maternal and infant death, we did not require continuous enrollment after delivery for the death outcome only. We defined eligibility criteria and covariates based on information available prior to or during pregnancy to minimize conditioning on variables potentially affected by the outcomes (**Supplement Figure 1**).

### Primary Exposure

Our exposure of interest was any prescription stimulant treatment during pregnancy. We used pharmacy claims to identify dispensing of amphetamine- and methylphenidate-class stimulants, excluding non-target agents such as modafinil, armodafinil, and pitolisant. For each prescription, we obtained fill dates and days’ supply. We defined pregnancy exposure as any stimulant prescription with days’ supply overlapping the interval from 14 days after LMP (approximate time of conception) through delivery, recognizing that some early first-trimester exposure may occur before pregnancy recognition. We characterized stimulant treatment as total days supplied during pregnancy, but in the primary analysis defined a binary exposure variable of any stimulant treatment in pregnancy versus none.

### Exposure to Opioid Use Disorder Medication

We used pharmacy dispensing claims to construct a person-level buprenorphine exposure history during pregnancy, excluding formulations indicated for pain. Because TennCare did not cover methadone for OUD treatment during the study period, buprenorphine was the primary OUD medication of interest. For each pregnancy, we calculated total buprenorphine days supplied from LMP through delivery and defined exposure as ≥14 days, based on a prior study with a similar population.^21^ Because buprenorphine treatment may function as a marker of OUD treatment engagement and lie on the pathway to some outcomes, it was not included in the primary propensity score model and examined separately in secondary analyses.

### Maternal Outcomes

The primary outcome was non-fatal opioid or non-opioid overdose occurring from delivery through one year postpartum, identified using diagnosis and procedure codes from inpatient, emergency department, and outpatient claims. Secondary outcomes included two composite outcomes (cardiovascular complications and severe maternal morbidity) and two individual outcomes (maternal death and buprenorphine treatment). We defined cardiovascular complications as a composite including chronic or gestational hypertension, preeclampsia, stroke or puerperal cerebrovascular events, aneurysm, cardiac rhythm disturbances, heart failure, cardiac arrest, and myocardial infarction, identified from birth through six weeks postpartum using birth certificates and claims. Severe maternal morbidity was defined according to Centers for Disease Control and Prevention criteria as the presence of at least one of 21 severe morbidity indicators or intensive care unit admission during the delivery hospitalization through six weeks postpartum.^22^ Maternal death was from any cause within one year postpartum. All outcomes were coded as binary indicators at the pregnancy level.

### Neonatal Outcomes

The primary outcome was preterm birth, defined as gestational age less than 37 completed weeks on the birth certificate. Secondary outcomes included low birth weight, defined as birth weight less than 2,500 grams on the birth certificate; NICU admission during the birth hospitalization, as recorded on the birth certificate and hospital claims; infant all-cause death within 365 days of life, identified from linked death certificates; and NOWS, ascertained from the birth hospitalization and any subsequent neonatal readmissions using ICD-9-CM and ICD-10-CM diagnosis codes on linked claims and hospital records up to 30 days after birth. All outcomes were binary indicators.

### Covariates

We selected covariates *a priori* based on clinical knowledge and prior literature regarding factors associated with stimulant prescribing and maternal outcomes.^12^ We ascertained covariates during pregnancy from LMP through delivery, before measurement of postpartum outcomes. From birth certificates, we obtained maternal age at delivery, race and ethnicity (categorized as non-Hispanic White, non-Hispanic Black, Hispanic, or other), educational attainment (categorized from less than high school through postgraduate and unknown), marital status (married vs not/unknown), and rural-urban residence using Rural-Urban Continuum Codes (2013). Reproductive and clinical characteristics included parity, pre-pregnancy body mass index, history of cesarean delivery, pre-pregnancy diabetes, gestational diabetes, number of prenatal visits, and birth year.

Smoking during pregnancy was derived from birth certificate data on cigarette and pack use and coded as a binary variable. From TennCare claims and linked data, we ascertained StUD, alcohol use disorder, cannabis use disorder, and mental health diagnoses during the pre-delivery ascertainment window, including depressive, anxiety, bipolar, psychotic, and related disorders. ADHD was identified from claims and included as its own binary covariate rather than grouped with the broader mental health diagnosis indicator. Sexually transmitted infections, including gonorrhea, chlamydia, and syphilis, were also included. The propensity score models used the same covariate set as presented in the baseline characteristics table plus birth year.

### Statistical Analysis

All analyses classified pregnancies according to any prescription stimulant exposure during pregnancy, regardless of subsequent discontinuation or dose changes. We summarized baseline characteristics overall and by stimulant exposure status using medians and interquartile range for continuous variables and counts and percentages for categorical variables; we used Wilcoxon rank-sum tests and χ^2^ or Fisher exact tests for group comparisons as appropriate.

We used the propensity score to calculate overlap weights to estimate associations between stimulant exposure and outcomes. Overlap weighting emphasizes the region of common support between treated and untreated groups, reduces variance relative to inverse probability weighting, and minimizes the influence of extreme propensity scores (**Supplement Figure 2**).^23^ We constructed a propensity score dataset using single imputation for limited missing covariate data, imputing missing maternal age using the analytic sample median, pre-pregnancy body mass index using the analytic sample mean, and prenatal visits as zero when absent from claims. We excluded pregnancies with missing categorical covariates. Logistic regression estimated the propensity of receiving stimulant treatment in pregnancy as a function of the covariates and birth year; overlap weights targeted the region of common support.^23^ The propensity score model included the full covariate set, including StUD and ADHD indicators, plus birth year. In secondary analyses, we then stratified the cohort by presence of StUD and by ADHD diagnosis and re⍰estimated the overlap⍰weighted associations within these subgroups.

For the primary analysis, we assessed outcomes among those with versus without stimulant use during pregnancy. Given that buprenorphine could be a potential mediator if stimulant use increases buprenorphine initiation and adherence or as a confounder if buprenorphine use increases the likelihood of initiating stimulant treatment, we included models with and without buprenorphine in the propensity score.^21,24^ In models including buprenorphine, we also assessed for potential effect measure modification by stimulant-by-buprenorphine interaction terms, to evaluate whether concurrent buprenorphine treatment strengthened or attenuated the association between stimulant exposure and maternal and neonatal outcomes, with statistical interaction evaluated using Wald tests. Furthermore, due to the potential for confounding by indication among those using prescription stimulants during pregnancy, we conducted secondary analyses stratified by ADHD and StUD, treating these subgroup analyses as exploratory. We used model-based marginal predictions to derive weighted outcome percentages and conducted all analyses in R version 4.3.3 (R Foundation for Statistical Computing, Vienna, Austria). Statistical significance was set at α = 0.05 for all tests, with 2-sided P values.

Because the clinical question is whether stimulant treatment adds meaningful risk rather than whether any risk difference exists, we prespecified interpretation based on the location and width of confidence intervals. Given the number of stimulant-exposed pregnancies and baseline outcome risks, the analysis was sized to detect odds ratios of approximately 1.9 for postpartum overdose and 1.5 for preterm birth with 80% power; confidence limits are therefore reported for every outcome so that the range of risk compatible with these data is explicit. Interaction and subgroup tests were not adjusted for multiplicity and are reported as exploratory.

## RESULTS

### Cohort Characteristics

Among 12,327 pregnancies with OUD, 380 (3.1%) received prescription stimulants during pregnancy (**Figure 1**). Individuals treated with prescription stimulants were slightly older (median 29 vs 27 years), more often non-Hispanic White (94.7% vs 91.8%) and married (31.3% vs 26.7%), and more likely to have at least some college or an associate degree or higher (45.5% vs 29.2%), with a median of 116 days of stimulant exposure during pregnancy (**Table 1**). Stimulant-treated individuals had lower prevalences of alcohol use disorder (42.1% vs 53.1%, *P* < .001), cannabis use disorder (4.5% vs 9.8%, *P* < .001), and chlamydia infections (2.1% vs 4.4%, *P* = .03), with similar prenatal care utilization (median 9 vs 9 visits). ADHD diagnoses were substantially more common among stimulant-treated individuals (48.7% vs 1.2%, *P* < .001), whereas stimulant use disorder was modestly less common (9.2% vs 11.6%, *P* = .15). In unadjusted analyses, overdose, cardiovascular complications, severe maternal morbidity, preterm birth, low birth weight, and NOWS were similar between stimulant-exposed and unexposed pregnancies. Maternal death occurred only in the non-stimulant group and NICU admission was more frequent among stimulant-exposed infants (**Table 2)**.

**Figure 1.**
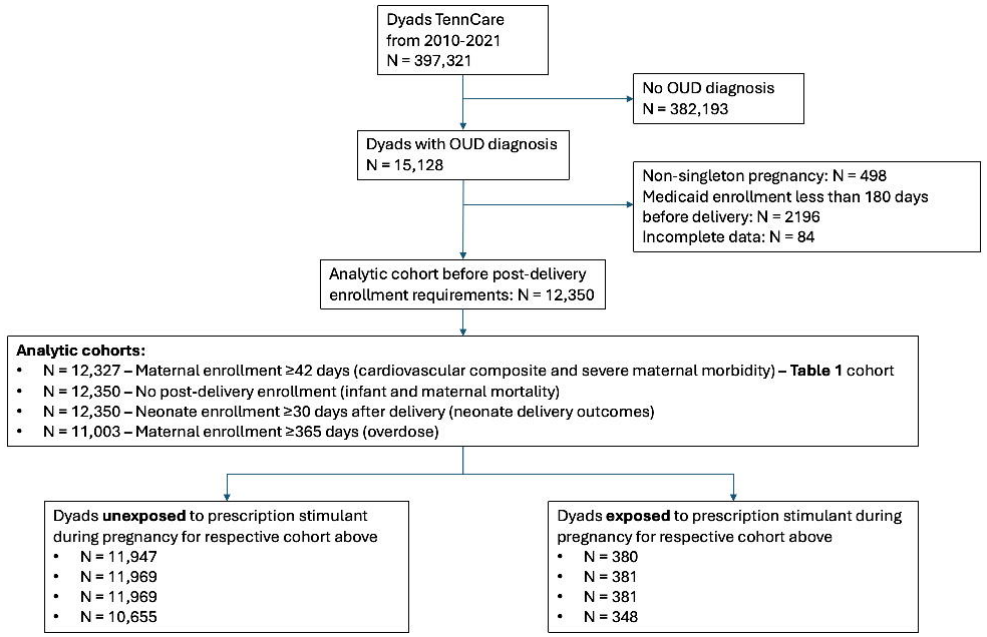
Cohort Selection Flow Diagram.

**Table 1.** Baseline Characteristics of Pregnant Individuals with OUD by Prescription Stimulant Exposure.

| **Characteristic** | **No prescription stimulant<br>(n = 11,947)** | **Prescription stimulant<br>(n = 380)** |
|---|---|---|
|  | **n (%)** | **n (%)** |
| **Demographics** |  |  |
| **Maternal age, median (IQR), years** | 27.0 (24.0-31.0) | 29.0 (25.0-33.0) |
| **Race and ethnicity** |  |  |
| Non-Hispanic White | 10965 (91.8) | 360 (94.7) |
| Non-Hispanic Black | 876 (7.3) | 14 (3.7) |
| Hispanic | 95 (0.8) | 6 (1.6) |
| Other | 11 (0.1) | 0 (0.0) |
| **Married** | 3187 (26.7) | 119 (31.3) |
| Unknown | 21 (0.2) | 0 (0.0) |
| **BMI, median (IQR)** | 23.2 (20.4-27.4) | 23.2 (21.1-26.7) |
| **Education** |  |  |
| 8<sup>th</sup> grade or less | 155 (1.3) | 3 (0.8) |
| 9<sup>th</sup>-12<sup>th</sup> but no diploma | 2628 (22.0) | 60 (15.8) |
| Highschool graduate or GED | 5623 (47.1) | 144 (37.9) |
| Some college but no degree | 2933 (24.6) | 130 (34.2) |
| Associate’s degree | 383 (3.2) | 33 (8.7) |
| Bachelor’s degree | 157 (1.3) | 7 (1.8) |
| Master’s degree | 17 (0.1) | 3 (0.8) |
| Doctorate or professional degree | 4 (0.0) | 0 (0.0) |
| Unknown | 47 (0.4) | 0 (0.0) |
| **Rurality** |  |  |
| > 1 million | 2907 (24.3) | 156 (41.1) |
| 250,000 - 1 million | 4177 (35.0) | 103 (27.1) |
| < 250,000 | 1304 (10.9) | 23 (6.1) |
| > 20,000, adjacent to metro area | 1236 (10.3) | 32 (8.4) |
| > 20,000, not adjacent to metro area | 59 (0.5) | 1 (0.3) |
| 5,000 - 20,000 adjacent to metro area | 1446 (12.1) | 39 (10.3) |
| 5,000 - 20,000 not adjacent to metro area | 368 (3.1) | 21 (5.5) |
| < 5,000 adjacent to a metro area | 265 (2.2) | 4 (1.4) |
| < 5,000 not adjacent to metro area | 185 (1.5) | 1 (0.3) |
| **Stimulant days in pregnancy** | 0 (0) | 116 (60-180) |
| **Mental health diagnoses** | 6573 (55.0) | 258 (67.9) |
| **ADHD** | 146 (1.2) | 185 (48.7) |
| **Prepregnancy diabetes** | 142 (1.2) | 3 (0.8) |
| **Gestational diabetes** | 646 (5.4) | 17 (4.5) |
| **Substance use** |  |  |
| Smoking | 8332 (69.7) | 242 (63.7) |
| Stimulant use disorder | 1385 (11.6) | 35 (9.2) |
| Alcohol use disorder | 6340 (53.1) | 160 (42.1) |
| Cannabis use disorder | 1174 (9.8) | 17 (4.5) |
| **Sexually transmitted infection** |  |  |
| Chlamydia | 521 (4.4) | 8 (2.1) |
| Gonorrhea | 112 (0.9) | 3 (0.8) |
| Syphilis | 34 (0.3) | 1 (0.3) |
| **Previous births, median (IQR)** | 1.0 (1.0-2.0) | 2.0 (1.0-3.0) |
| **Previous cesarean delivery** | 2408 (20.2) | 89 (23.4) |
| **Prenatal visits, median (IQR)** | 9.0 (5.0-12.0) | 9.0 (5.0-12.0) |

Abbreviations: OUD, opioid use disorder; IQR, interquartile range; ADHD, attention-deficit/hyperactivity disorder; GED, general educational development.

**Table 2.** Unadjusted Frequencies and Adjusted Associations of Prescription Stimulant Exposure With Maternal and Neonatal Outcomes.

|  | **No stimulant<br>(n = 11,947)** | **Stimulant<br>(n = 380)** |  |  |
|---|---|---|---|---|
|  | **No. (%)** | **No. (%)** | **aOR (95% CI)** | ***P* value** |
| **Maternal outcomes** |  |  |  |  |
| Overdose (primary) | 405 (3.4) | 16 (4.2) | 1.13 (0.61-2.08) | .70 |
| Cardiovascular complications | 1160 (9.7) | 44 (11.6) | 1.17 (0.81-1.70) | .40 |
| Severe maternal morbidity | 713 (6.0) | 21 (5.5) | 1.02 (0.61-1.69) | .94 |
| Death | 62 (0.5) | 0 (0.0) | - | - |
| Buprenorphine treatment | 5989 (50.1) | 192 (50.5) | 1.09 (0.85-1.39) | .51 |
| **Neonatal outcomes** | **n = 11,969** | **n = 381** |  |  |
| Preterm birth (primary) | 1728 (14.4) | 56 (14.7) | 1.04 (0.74-1.46) | .81 |
| Low birth weight | 2021 (16.9) | 52 (13.6) | 0.77 (0.55-1.10) | .15 |
| NICU | 1727 (14.4) | 74 (19.4) | 1.58 (1.17-2.14) | .003 |
| Death | 111 (0.9) | 5 (1.3) | 2.27 (0.86-5.99) | .10 |
| NOWS | 5371 (44.9) | 169 (44.4) | 1.25 (0.99-1.59) | .06 |

aORs (adjusted Odds Ratio) from overlap-weighted logistic regression models; reference group is pregnancies without prescription stimulant exposure. Adjusted outcomes from cohorts shown in **Figure 1**.

### Maternal outcomes

In adjusted analyses, prescription stimulant treatment was not associated with different odds of postpartum overdose (adjusted odds ratio [aOR], 1.13; 95% CI, 0.61–2.08), and adjusted predicted risks were similar for stimulant-treated individuals and those not treated (approximately 4%) (**Table 2; Figure 2)**. Prescription stimulant exposure was also not associated with increased cardiovascular complications (aOR, 1.17; 95% CI, 0.81–1.70) or severe maternal morbidity (aOR, 1.02; 95% CI, 0.61–1.69), with comparable adjusted predicted risks between groups. Maternal death was rare and occurred only among individuals without prescription stimulant exposure. Stimulant treatment was not associated with buprenorphine treatment during pregnancy (aOR, 1.09; 95% CI, 0.85–1.39).

**Figure 2.**
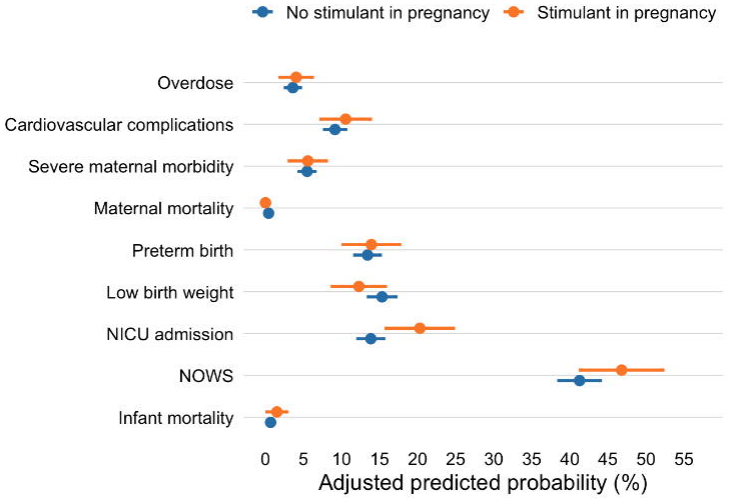
Probability of Maternal and Neonatal Outcomes by Prescription Stimulant Exposure.

In models additionally adjusted for buprenorphine treatment, the association between stimulant treatment and overdose remained null (aOR, 1.13; 95% CI, 0.61–2.09; **Supplement Table 1**). In exploratory models including stimulant-by-buprenorphine interaction terms, the interaction for overdose was statistically significant (P for interaction = .03). Relative to pregnancies exposed to neither medication, odds of overdose were 1.82 (95% CI, 1.00–3.33) among pregnancies receiving both stimulants and buprenorphine, corresponding to predicted risks of 6.7% (95% CI, 3.8–11.4) versus 3.8% (95% CI, 3.3–4.3); the stimulant-only stratum was below the reference (aOR, 0.62; 95% CI, 0.23–1.70). Interactions for all other outcomes were not significant (P ≥ .15). Subgroup analyses largely mirrored the main maternal results, apart from higher cardiovascular risk among individuals with ADHD receiving stimulants (aOR, 2.33; 95% CI, 1.13–4.82; **Supplement Table 2**).

### Neonatal outcomes

In adjusted analyses, prescription stimulant treatment during pregnancy was not associated with different odds of preterm birth (aOR, 1.04; 95% CI, 0.74–1.46), and adjusted predicted risks of preterm birth were similar for stimulant-exposed and unexposed infants (approximately 13%) (**Table 2; Figure 2**). Low birth weight, infant death through 1 year, and NOWS were also not associated with prescription stimulant treatment. In contrast, stimulant exposure was associated with higher odds of NICU admission (aOR, 1.58; 95% CI, 1.17– 2.14), with adjusted predicted probabilities of 13.8% among infants unexposed to stimulants and 20.3% among stimulant-exposed neonates.

In models additionally adjusted for buprenorphine, estimates for preterm birth, NICU admission, and NOWS were nearly identical to the main results (aOR [95% CI] for preterm birth, 1.05 [0.75–1.47]; NICU admission, 1.59 [1.18–2.15]; NOWS, 1.25 [0.99–1.59]), and predicted NICU risks remained approximately 14% vs 20% by stimulant exposure (**Supplement Table 1**). Interaction and subgroup analyses for preterm birth, NICU admission, and NOWS were imprecise but generally supported the primary findings, with no evidence of additional risk from stimulant treatment, including in analyses restricted to individuals with documented StUD or ADHD (**Supplement Tables 1 and 2**).

## DISCUSSION

In this statewide cohort of pregnant individuals with OUD, prescription stimulant treatment during pregnancy was not associated with increased risk of postpartum overdose, cardiovascular complications, severe maternal morbidity, maternal death, preterm birth, low birth weight, or NOWS. In contrast, stimulant exposure was associated with modestly higher odds of NICU admission. These findings suggest that, within the context of contemporary perinatal OUD care, clinically indicated prescription stimulant treatment does not appear to confer large additional maternal or infant safety risk but may be associated with greater NICU utilization.

The maternal findings align with prior literature suggesting that prescribed psychostimulants, when used in monitored settings, do not substantially increase serious maternal complications in pregnancy.^14,15,17^ Despite longstanding concern about the vasoconstrictive and sympathomimetic effects of stimulants, stimulant treatment in this cohort was not associated with excess cardiovascular complications or severe maternal morbidity.^11,12^ Earlier work in pregnant individuals with OUD linked prescription psychostimulant use to improved buprenorphine initiation and retention and fewer substance use disorder-related admissions.^21^ Our findings add no clear evidence of increased maternal harm. The current findings support a harm-reduction framework in which carefully monitored prescription stimulant treatment can be integrated into perinatal OUD care without observable increases in major maternal safety events.^6,19,24^

Large ADHD-focused studies in general obstetric populations have not found strong associations with major congenital anomalies or neurodevelopmental disorders, though some report modest increases in preterm birth.^14–18^ In contrast, among pregnancies defined by OUD, where baseline vulnerability and polysubstance exposure are high, prenatal stimulant treatment was not associated with higher risks of preterm birth or low birth weight; estimates for infant death were near the null but imprecise because events were uncommon. The higher NICU admission risk observed among stimulant-exposed neonates may reflect combined effects of stimulant-related neuroautonomic activation, heightened clinical concern about polysubstance exposure, and structured monitoring protocols for opioid-exposed infants that can lower thresholds for intensive care. Consistent with a surveillance rather than a morbidity explanation, stimulant exposure was not associated with preterm birth, low birth weight, or mean gestational age, the clinical drivers that ordinarily accompany a genuine increase in neonatal acuity.^25,26^ Although the odds of NOWS were higher without reaching statistical significance among stimulant-exposed infants, absolute NOWS risks were high in both groups after accounting for buprenorphine treatment, suggesting that background opioid exposure and OUD care pathways may be drivers of withdrawal severity.^4,5,25–28^ These patterns suggest that NICU admission in this setting may often reflect conservative surveillance within integrated OUD care rather than severe or irreversible morbidity.^4,6,29^

Because stimulant and buprenorphine exposures overlapped in time, stratified and interaction analyses are best interpreted as exploratory. In interaction models, odds of overdose were highest among pregnancies receiving both medications and the interaction term reached nominal significance. We regard this as hypothesis-generating rather than causal for three reasons. First, the lower confidence limit abuts the null and few events occurred within the combined stratum. Second, the pattern is difficult to reconcile pharmacologically: overdose odds in the buprenorphine-only stratum were below the unexposed reference, and the stimulant-only stratum was also below reference, so the combined stratum is more plausibly capturing pregnancies with greater underlying substance use severity and more frequent health care contact than an adverse drug interaction. Third, interaction terms were tested across six outcomes without adjustment for multiplicity. The same caution applies to the elevated cardiovascular composite among individuals with ADHD receiving stimulants, which rests on a subgroup of 331 pregnancies and was not accompanied by any difference in severe maternal morbidity. Both findings are priorities for replication in larger multi-state cohorts rather than evidence of harm. Together, these patterns suggest that receipt of prescription stimulants may mark differences in treatment engagement, clinical stability, or provider decision-making, rather than directly causing differences in overdose or neonatal outcomes, consistent with the “healthy user” effect described in pharmacoepidemiology.^21,30^

These findings have implications for addiction treatment delivery and coverage policy. Obstetric clinicians, addiction medicine specialists, and payers are often reluctant to continue or initiate controlled substances, including stimulants, in pregnant individuals with OUD, and coverage for stimulant medications in Medicaid programs is frequently restrictive.^12,19^ At the same time, no FDA-approved pharmacotherapy exists for StUD and current guidelines identify contingency management as the standard of care while acknowledging off-label pharmacotherapy use.^19^ Our findings, combined with prior evidence of improved engagement in OUD medication treatment, suggest that clinically indicated prescription stimulants may be continued during pregnancy, guided by individualized risk-benefit assessment rather than routine discontinuation based on pregnancy or OUD diagnosis alone.^21^ Shared decision-making remains essential, recognizing that stimulants are not an approved treatment for StUD and that evidence for their use in this context remains limited.^7–9,19^

### Limitations

This observational analysis of administrative claims is subject to residual confounding from unmeasured factors, including substance use severity, treatment motivation, social drivers of health, and provider-level influences on prescribing and outcomes. Pharmacy claims capture dispensing rather than ingestion, and our exposure definition based on any stimulant prescription overlapping pregnancy does not incorporate dose, timing, or duration beyond days supplied. Although the overall cohort was large, rare outcomes such as maternal and infant death yielded few events, limiting statistical power and producing imprecise estimates, and exploratory interaction and subgroup analyses were constrained by small stimulant-exposed strata, increasing potential for instability and overinterpretation. Information on long-term child neurodevelopmental outcomes was not available. Finally, we examined multiple maternal and neonatal outcomes and multiple interaction and subgroup terms without formal adjustment for multiplicity, so nominal P values should be read accordingly, and the two positive exploratory findings should not be treated as confirmatory.

These findings derive from a Medicaid-insured population in Tennessee and may be most generalizable to pregnant individuals with OUD receiving care in similar state Medicaid programs and integrated perinatal addiction services. Results may not fully extend to commercially insured populations, uninsured individuals, or settings with different OUD treatment access, stimulant prescribing practices, or patterns of polysubstance use. Because TennCare did not cover methadone for OUD during the study period, findings also may not generalize to systems in which methadone is the predominant medication for OUD. Nonetheless, the large statewide cohort and linkage of claims with vital records support external validity for comparable health systems

## CONCLUSION

Among pregnant individuals with OUD, prenatal prescription stimulant treatment was not associated with increased odds of postpartum overdose, cardiovascular complications, severe maternal morbidity, preterm birth, low birth weight, or NOWS. Stimulant-exposed infants were more likely to be admitted to the NICU without corresponding differences in gestational age, birth weight, or withdrawal, a pattern more consistent with heightened surveillance than with neonatal harm. These first OUD-specific estimates do not support routine discontinuation of clinically indicated stimulant therapy, and they argue against coverage policies that restrict access on the basis of pregnancy or OUD diagnosis alone. Two exploratory signals, elevated overdose odds with concurrent buprenorphine and an elevated cardiovascular composite within the ADHD subgroup, require replication before they can inform practice.

## Supporting information

Supplement Figure 1. Timing of Covariate, Exposure, and Outcome Ascertainment Relative to Delivery.

Supplement Figure 2. Covariate Balance Before and After Overlap Weighting.

## Data Availability

Data are not available.

## Acknowledgements

Tennessee Department of Health

## Declaration of generative AI and AI-assisted technologies in the manuscript preparation process

During the preparation of this work, the author(s) used Claude for refining the text. The author(s) reviewed and edited the output as needed and take full responsibility for the content of the published article

---

**Supplement Figure 1. Timing of Covariate, Exposure, and Outcome Ascertainment Relative to Delivery**.

**Supplement Figure 2. Covariate Balance Before and After Overlap Weighting**.

