## Supplement Tables for "Maternal and Neonatal Outcomes Associated with Prenatal Prescription Stimulant Treatment Among Pregnant Individuals with Opioid Use Disorder"

| Supplement Table 1. Association of Prescription Stimulant Exposure with Maternal and Neonatal Outcomes: Buprenorphine-Adjusted and Interaction Models Among Pregnant Individuals with OUD. | | | | | |
| --- | --- | --- | --- | --- | --- |
| Model | | | **Comparison** | **aOR (95% CI)** | **Predicted probability,**  **% (95% CI)** |
| Overdose | | |  |  |  |
|  | Direct effect (buprenorphine adjusted) | | No stimulant vs Stimulant | 1.13 (0.61-2.09) | 3.6 (2.6-4.6) vs 4.0 (1.9-6.1) |
|  | Interaction model | | No stimulant & No buprenorphine | 1 [Reference] | 3.8 (3.3-4.3) |
|  |  |  | Stimulant & No buprenorphine | 0.62 (0.23–1.70) | 2.4 (0.9–6.2) |
|  |  |  | No stimulant & buprenorphine | 0.80 (0.65–0.99) | 3.0 (2.6–3.5) |
|  |  |  | Stimulant & buprenorphine | 1.82 (1.00–3.33) | 6.7 (3.8–11.4) |
|  | P for interaction = .03 | |  |  |  |
| Cardiovascular composite | | |  |  |  |
|  | Direct effect (buprenorphine adjusted) | | No stimulant vs Stimulant | 1.18 (0.81-1.70) | 9.1 (7.7-10.5) vs 10.5 (7.3-13.8) |
|  | Interaction model | | No stimulant & No buprenorphine | 1 [Reference] | 11.2 (10.4–12.0) |
|  |  |  | Stimulant & No buprenorphine | 1.27 (0.83–1.94) | 13.8 (9.6–19.5) |
|  |  |  | No stimulant & buprenorphine | 0.71 (0.63–0.80) | 8.2 (7.5–8.9) |
|  |  |  | Stimulant & buprenorphine | 0.82 (0.50–1.34) | 9.4 (6.0–14.4) |
|  | P for interaction = .77 | |  |  |  |
| Severe maternal morbidity | | |  |  |  |
|  | Direct effect (buprenorphine adjusted) | | No stimulant vs Stimulant | 1.02 (0.61-1.70) | 5.4 (4.4-6.5) vs 5.6 (3.1-8.0) |
|  | Interaction model | | No stimulant & No buprenorphine | 1 [Reference] | 6.9 (6.3–7.6) |
|  |  |  | Stimulant & No buprenorphine | 0.68 (0.34–1.33) | 4.8 (2.5–8.9) |
|  |  |  | No stimulant & buprenorphine | 0.71 (0.61–0.83) | 5.0 (4.5–5.6) |
|  |  |  | Stimulant & buprenorphine | 0.90 (0.50–1.62) | 6.3 (3.6–10.7) |
|  | P for interaction = .18 | |  |  |  |
| Preterm birth | | |  |  |  |
|  | Direct effect (buprenorphine adjusted) | | No stimulant vs Stimulant | 1.05 (0.75-1.47) | 13.3 (11.7-15.0) vs 13.9 (10.2-17.6) |
|  | Interaction model | | No stimulant & No buprenorphine | 1 [Reference] | 17.3 (16.3–18.2) |
|  |  |  | Stimulant & No buprenorphine | 0.98 (0.66–1.44) | 16.9 (12.2–23.0) |
|  |  |  | No stimulant & buprenorphine | 0.63 (0.57–0.70) | 11.6 (10.8–12.5) |
|  |  |  | Stimulant & buprenorphine | 0.69 (0.44–1.06) | 12.5 (8.5–18.0) |
|  | P for interaction = .73 | |  |  |  |
| NICU | | |  |  |  |
|  | Direct effect (buprenorphine adjusted) | | No stimulant vs Stimulant | 1.59 (1.18-2.15) | 13.8 (12.1-15.5) vs 20.3 (15.9-24.7) |
|  | Interaction model | | No stimulant & No buprenorphine | 1 [Reference] | 15.2 (14.3–16.2) |
|  |  |  | Stimulant & No buprenorphine | 1.54 (1.08–2.20) | 21.7 (16.4–28.1) |
|  |  |  | No stimulant & buprenorphine | 0.88 (0.79–1.0) | 13.6 (12.8–14.5) |
|  |  |  | Stimulant & buprenorphine | 1.16 (0.79–1.69) | 17.2 (12.5–23.2) |
|  | P for interaction = .55 | |  |  |  |
| NOWS | | |  |  |  |
|  | Direct effect (buprenorphine adjusted) | | No stimulant vs Stimulant | 1.25 (0.99-1.59) | 41.3 (38.5-44.0) vs 46.8 (41.4-52.2) |
|  | Interaction model | | No stimulant & No buprenorphine | 1 [Reference] | 34.5 (33.3–35.7) |
|  |  |  | Stimulant & No buprenorphine | 1.14 (0.85–1.54) | 37.6 (31.0–44.7) |
|  |  |  | No stimulant & buprenorphine | 2.34 (2.18–2.52) | 55.2 (54.0–56.5) |
|  |  |  | Stimulant & buprenorphine | 1.98 (1.49–2.64) | 51.0 (44.0–58.0) |
|  | P for interaction = .15 | |  |  |  |

Abbreviations: aOR, Adjusted Odds Ratio.

| Supplement Table 2. Sensitivity Adjusted Analyses Stratified by StUD (No stimulant = 1,385; Stimulant = 35) and ADHD (No stimulant = 146; Stimulant = 185). | | | | | |
| --- | --- | --- | --- | --- | --- |
| Outcome | | **aOR (95% CI)** | ***P* value** | **Predicted probability, % (95% CI)** | |
|  | |  |  | **No stimulant** | **Stimulant** |
| Maternal | |  |  |  |  |
| StUD | | |  |  |  |
|  | Overdose | 0.79 (0.22–2.79) | .72 | 10.0 (4.6–15.4) | 8.1 (-0.4–16.6) |
|  | Cardiovascular composite | 0.76 (0.22–2.58) | .65 | 8.3 (4.5–12.2) | 6.4 (-1.0–13.8) |
|  | Severe maternal morbidity | - | - | 6.6 (3.2-10.0) | 0.0 (0.0-0.0) |
| ADHD | | |  |  |  |
|  | Overdose | 1.19 (0.45–3.18) | .72 | 4.6 (1.4–7.8) | 5.4 (2.0–8.9) |
|  | Cardiovascular composite | 2.33 (1.13–4.82) | .02 | 8.1 (3.5–12.6) | 17.0 (11.0–23.0) |
|  | Severe maternal morbidity | 0.96 (0.32–2.82) | .94 | 4.1 (1.0–7.3) | 4.0 (1.0–6.9) |
| Neonate | |  |  |  |  |
| StUD | | |  |  |  |
|  | Preterm birth | 1.09 (0.40–2.98) | .87 | 17.4 (11.7–23.1) | 18.6 (6.5–30.8) |
|  | NICU | 1.99 (0.83–4.76) | .12 | 19.4 (13.0–25.8) | 32.4 (17.0–47.8) |
|  | NOWS | 1.48 (0.70–3.13) | .31 | 48.4 (39.1–57.7) | 58.1 (42.1–74.1) |
| ADHD | | |  |  |  |
|  | Preterm birth | 1.81 (0.91–3.60) | .09 | 9.4 (4.5–14.4) | 15.9 (10.5–21.3) |
|  | NICU | 1.40 (0.71–2.77) | .33 | 11.6 (6.1–17.0) | 15.5 (9.9–21.0) |
|  | NOWS | 1.13 (0.71–1.78) | .61 | 36.1 (28.1–44.0) | 38.9 (31.3–46.5) |

Abbreviations: aOR, Adjusted Odds Ratio; OUD, opioid use disorder; StUD, stimulant use disorder; ADHD, attention-deficit/hyperactivity disorder. **Note:** Maternal mortality, infant mortality, and low birthweight were too rare in these restricted cohorts to yield stable estimates and are therefore not presented.

| **Supplement Table 3. Gestational Age by Stimulant Exposure.** | | | | |
| --- | --- | --- | --- | --- |
| **Outcome** | **Model** | **Exposure contrast** | **Mean difference in weeks (95% CI)** | ***P* value** |
| Gestational age (weeks) | Total-effect | Stimulant vs no stimulant | 0.02 (-0.19-0.23) | .83 |
| Gestational age (weeks) | MOUD-adjusted | Stimulant vs no stimulant | 0.02 (-0.19-0.23) | .84 |
