## Supplement Figure 1. Timing of Covariate, Exposure, and Outcome Ascertainment Relative to Delivery. for "Maternal and Neonatal Outcomes Associated with Prenatal Prescription Stimulant Treatment Among Pregnant Individuals with Opioid Use Disorder"

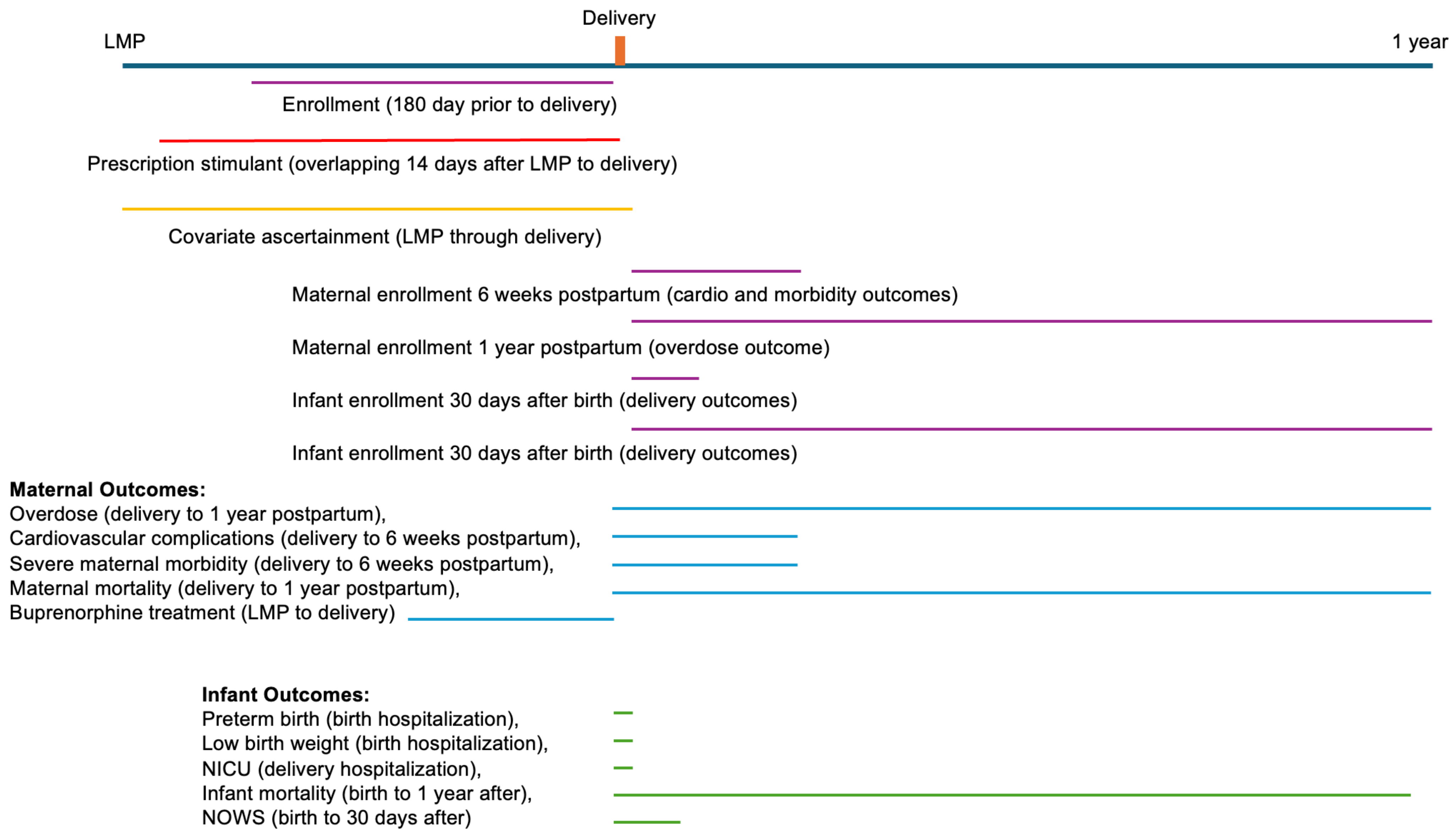
Delivery
LMP
1 year
Enrollment (180 day prior to delivery)
Prescription stimulant (overlapping 14 days after LMP to delivery)
Covariate ascertainment (LMP through delivery)
Maternal enrollment 6 weeks postpartum (cardio and morbidity outcomes)
Maternal enrollment 1 year postpartum (overdose outcome)
Infant enrollment 30 days after birth (delivery outcomes)
Infant enrollment 30 days after birth (delivery outcomes)
Maternal Outcomes:
Overdose (delivery to 1 year postpartum),
Cardiovascular complications (delivery to 6 weeks postpartum),
Severe maternal morbidity (delivery to 6 weeks postpartum),
Maternal mortality (delivery to 1 year postpartum),
Buprenorphine treatment (LMP to delivery)
Infant Outcomes:
Preterm birth (birth hospitalization),
Low birth weight (birth hospitalization),
NICU (delivery hospitalization),
Infant mortality (birth to 1 year after),
NOWS (birth to 30 days after)
