## Supplement Figure 2. Covariate Balance Before and After Overlap Weighting. for "Maternal and Neonatal Outcomes Associated with Prenatal Prescription Stimulant Treatment Among Pregnant Individuals with Opioid Use Disorder"

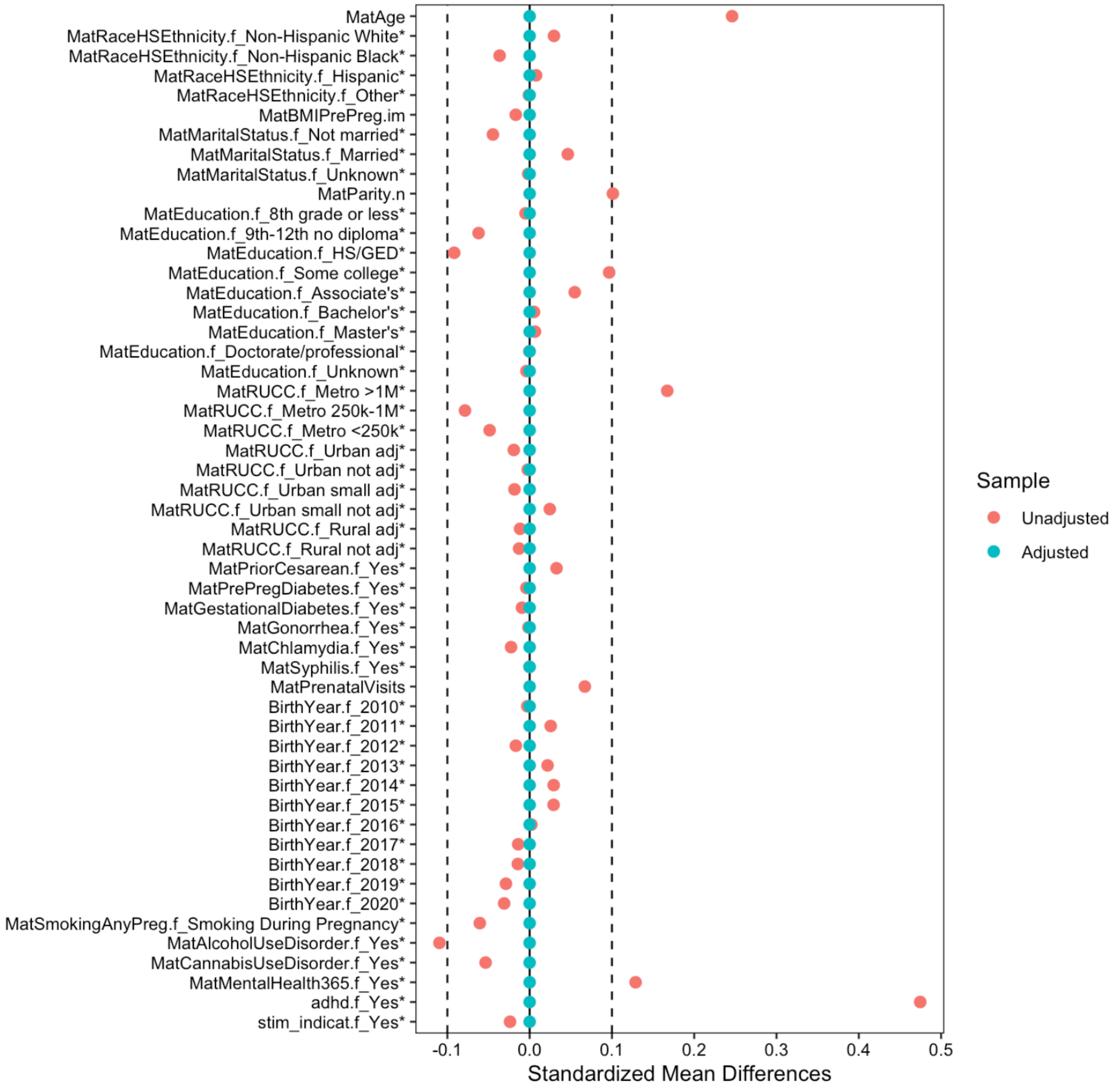
MatAge
MatRaceHSEthnicity.f_Non-Hispanic White*
MatRaceHSEthnicity.f_Non-Hispanic Black*
MatRaceHSEthnicity.f_Hispanic*
MatRaceHSEthnicity.f_Other*
MatBMIPrePreg.im
MatMaritalStatus.f_Not married*
MatMaritalStatus.f_Married*
MatMaritalStatus.f_Unknown*
MatParity.n
MatEducation.f_8th grade or less*
MatEducation.f_9th-12th no diploma*
MatEducation.f_HS/GED*
MatEducation.f_Some college*
MatEducation.f_Associate's*
MatEducation.f_Bachelor's*
MatEducation.f_Master's*
MatEducation.f_Doctorate/professional*
MatEducation.f_Unknown*
MatRUCC.f_Metro >1M*
MatRUCC.f_Metro 250k-1M*
MatRUCC.f_Metro <250k*
MatRUCC.f_Urban adj*
MatRUCC.f_Urban not adj*
MatRUCC.f_Urban small adj*
MatRUCC.f_Urban small not adj*
MatRUCC.f_Rural adj*
MatRUCC.f_Rural not adj*
MatPriorCesarean.f_Yes*
MatPrePregDiabetes.f_Yes*
MatGestationalDiabetes.f_Yes*
MatGonorrhea.f_Yes*
MatChlamydia.f_Yes*
MatSyphilis.f_Yes*
MatPrenatalVisits
BirthYear.f_2010*
BirthYear.f_2011*
BirthYear.f_2012*
BirthYear.f_2013*
BirthYear.f_2014*
BirthYear.f_2015*
BirthYear.f_2016*
BirthYear.f_2017*
BirthYear.f_2018*
BirthYear.f_2019*
BirthYear.f_2020*
MatSmokingAnyPreg.f_Smoking During Pregnancy*
MatAlcoholUseDisorder.f_Yes*
MatCannabisUseDisorder.f_Yes*
MatMentalHealth365.f_Yes*
adhd.f_Yes*
stim_indicat.f_Yes*
-0.1
0.0
0.1
0.2
0.3
0.4
0.5
Standardized Mean Differences
Sample
Unadjusted
Adjusted
